# The impact of occupational risk from COVID on GP supply in England: A cross-sectional study

**DOI:** 10.1101/2020.06.04.20122119

**Authors:** Miqdad Asaria, Rebecca Fisher

## Abstract

**Objectives:** To identify the risk of general practitioner mortality from COVID and the impact of measures to mitigate this risk on the level and socioeconomic distribution of primary care provision in the English NHS

**Design:** Cross sectional study

**Setting:** All GP practices providing primary care under the NHS in England

**Participants:** 45,858 GPs and 6,771 GP practices in the English NHS

**Main outcome measures:** Numbers of high-risk GPs, high-risk single-handed GP practices, patients associated with these high-risk single-handed practices and the regional and socioeconomic distribution of each. Mortality rates from COVID by age, sex and ethnicity were used to attribute risk to GPs and the Index of Multiple Deprivation was used to determine socioeconomic distributions of the outcomes.

**Results:** Of 45,858 GPs in our sample 3,632 (7.9%) were classified as high risk or very high risk. Of 6,771 GP practices in our sample 639 (9.4%) were identified as single-handed practices and of these 209 (32.7%) were run by a GP at high or very high risk. These 209 single-handed practices care for 710,043 patients. GPs at the highest levels of risk from COVID, and single-handed practices run by high-risk GPs were concentrated in the most deprived neighbourhoods in the country. London had the highest proportion of both GPs and single-handed GP practices at very high risk of COVID mortality with 1,160 patients per 100,000 population registered to these practices.

**Conclusions:** A significant proportion of GPs working in England, particularly those serving patients in the most deprived neighbourhoods, are at high risk of dying from COVID. Many of these GPs run single-handed practices. These GPs are particularly concentrated in London. There is an opportunity to provide additional support to mitigate COVID risk for GPs, GP practices and their patients. Failure to do so will likely exacerbate existing health inequalities.

**What is already known:**

- Known risk factors for morbidity and mortality from COVID-19 include age, sex, ethnicity and certain underlying health conditions.
- NHS England have suggested that NHS staff who may be at higher risk from COVID are risk assessed and have their activities adjusted accordingly, including ceasing face to face patient contact.

**What this study adds:**

- This study applies risk scoring to calculate the number of GPs practicing in England who are likely to be at high or very high risk of death from COVID. We examine the potential effect of removing GPs at high or very high risk from COVID from face to face patient contacts, estimating the number of GPs and patients likely to be affected, and relating this to deprivation and geography.
- We estimate that of 45,858 GPs in our sample, 2,253 (4.9%) were classified as high risk, and 1,379 (3%) as very high risk from COVID. These are likely to be conservative estimates.
- GPs at high risk of COVID are more likely to work in areas of high socioeconomic deprivation.
- Almost one in three single-handed GP practices (32.7%, or 209 out of 639) is run by a GP we estimate to be at high or very high risk from COVID. If these GPs did not see patients face to face, 710,043 patients would be left without face to face GP appointments. Single-handed GP practices in areas of high socioeconomic deprivation are more likely to be run by GPs at higher risk of COVID.

## Introduction

As the NHS shifts to the ‘second phase’ of responding to the COVID-19 (‘COVID’) pandemic, learning how to live and work alongside COVID is necessary. In general practice this will be challenging. Strategies already in use are likely to be maintained, including using ‘hot hubs’, ‘zoning’ and using telephone and video consulting to reduce face to face contact where possible. But these can only go so far. The number of patients requiring face to face consultations is likely to creep up over time (as examinations and tests can no longer be deferred), and with it the exposure of general practitioners (GPs) to COVID.

The risk of catching COVID – and of dying from it – is not equally distributed amongst GPs. Relatively early in the pandemic, Public Health England issued guidance identifying three risk factors used to guide managers in conversations with staff about increased vulnerability to COVID: age > 70 years, selected underlying health conditions, and pregnancy.(1) Although ethnicity was not originally included as a risk factor this has since been recognised as an important omission. Morbidity and mortality from COVID is higher in Black, Asian and Minority Ethnic (BAME) people than in Caucasian people, and the vast majority of COVID deaths in healthcare workers have been BAME staff, despite BAME workers accounting for 21% of the NHS workforce.(2–4)

The NHS Risk Reduction Framework attempts to capture this differential risk, with suggestion made – since reiterated in a letter from NHS England – that NHS employees deemed to be at higher risk from COVID be re-deployed to roles without face to face contact.(5,6) Unlike in secondary care, where individuals often work as part of large teams, GPs tend to work in smaller teams, and sometimes as the sole medical practitioner in a surgery (so called ‘single handed’ practice). The impact of removing GPs from face to face patient duties may be harder to compensate for, and in some cases may leave an entire patient population without a GP with whom they can face to face consult in a manner that is safe for GP and patient.

This analysis seeks to understand the potential implications of applying COVID-related occupational risk assessment to GPs. We calculate how many GPs currently practicing in England are likely to fall within high risk groups, whether those GPs are concentrated in particular geographical areas, and the correlation of this with socioeconomic deprivation – a known predictor of GP workload. We also calculate the number of single-handed practices being run by GPs likely to be deemed at high risk from COVID, and the number of patients covered by these GPs.

## Methods

### Workforce data

We used Primary care workforce data from the most recent release of the General Practice Workforce series published by NHS Digital for the 31^st^ March 2020.(7) This dataset includes details on all NHS GPs as well as all NHS GP Practices operating in England. Data on individuals in this data series was used to categorise GPs by age, sex, country of qualification, primary job role and clinical commissioning group. The data series on GP Practices was used to identify single-handed GP practices as those run by a single GP (excluding locums, registrars and retainers who may work at the practice). GPs and practices lacking data on GP age and sex were excluded as meaningful judgements on risk of COVID mortality could not be made without these key characteristics. This resulted in 1,615 (3.4%) of GPs and 4 (0.06%) of single-handed GP practices being excluded from our dataset.

### Age-specific COVID mortality rates

Age and sex specific mortality data covering the period between the 1^st^ March 2020 and 30^th^ April 2020 from the Office of National Statistics (ONS) were used to capture deaths from COVID.(8) This mortality data was combined with age and sex specific population data from ONS mid-year population estimates for 2019 to calculate age-sex specific COVID mortality rates.(9)

### Socioeconomic distribution of Clinical Commissioning Groups

The Index of Multiple Deprivation (IMD) 2019 measures relative deprivation in small areas in England called lower-layer super output areas (LSOAs).(10) We attributed IMD scores from LSOAs to CCGs by calculating population weighted averages, and used these attributed scores to rank CCGs into population weighted deprivation quintile groups with each quintile group of CCGs covering approximately a fifth of the total population.(11–13) A table detailing the CCG deprivation quintiles we calculated for use in this analysis is provided in the supplementary appendix (Table A2).

### COVID risk reduction framework

We used the Risk Reduction Framework for NHS Staff at risk of COVID infection to guide our understanding of key characteristics that contribute to the level of risk from COVID faced by GPs.(5) The framework highlights age, sex, ethnicity, underlying health conditions and pregnancy as the five most important factors that influence risk from COVID among NHS staff. We also considered the Safety Assessment And Decision (SAAD) score designed to assess risks for BAME communities during a COVID pandemic infection in general practice which highlights similar factors as contributing to mortality risk using a more detailed treatment of ethnicity and underlying health conditions.(14) We used data on age, sex and ethnicity to characterise risk amongst GPs using the age-groups and ethnicity categories suggested in the framework. Data on underlying health conditions and pregnancy were not available in the workforce data so we were unable to include these in our analysis. Additionally, ethnicity data was not provided in the data so we used country of qualification as a proxy for ethnicity assuming that GPs listed as qualifying either in the United Kingdom (UK) or the European Economic Area (EEA) as being White whilst GPs listed as qualifying anywhere else were characterised as BAME. Our ethnicity variable therefore mis-categorises those BAME GPs that qualified in the UK or EEA as White and hence will under-estimate COVID risk associated with these GPs. We have requested bespoke datasets from NHS Digital and the General Medical Council that explicitly include ethnicity and will update the analysis in this manuscript with more accurate ethnicity data if we are able to obtain these data.

### COVID risk categorisation of GPs

We used the age-sex specific COVID mortality rates we calculated to translate the risk reduction framework into a risk scoring system. Risk scores were calculated by dividing all mortality rates by those for women aged 55–60 (the lowest risk group for whom COVID mortality risk is non-negligible). These scores were adjusted for ethnicity using a recent ONS study that found that age and sex adjusted COVID mortality rates for non-White ethnic groups are between 2 times and 4 times those of Whites depending on which non-White ethnic group is being compared.(15) Two alternative sets of risk scores were calculated for BAME GPs by using the upper and lower range of these ethnicity specific adjustments respectively to adjust the overall age-sex specific risk scores. We used the resulting risk scores to categorise both GPs and single-handed GP practices into 4 risk categories (Low, Medium, High and Very High) to reflect risk of mortality from COVID.

### Outcome measures

Our main outcome measures were (1) numbers of GPs, (2) number of single-handed GP practices and (3) numbers of patients registered to single-handed GP practices. Each outcome measure was disaggregated by COVID risk category to identify the numbers of GPs, single-handed practices and patients registered to single-handed practices at high and very high risk levels. The regional and socioeconomic distribution of high and very high COVID risk in primary care was examined for each outcome measure.(16)

### Software

All analysis was conducted using R version 4.0.0 statistical software.(17)

## Results

Age-sex specific COVID mortality rates show that risk increases rapidly with age, with risk of dying from COVID in those over 70 years of age being approximately a hundred times higher than risk for those under 55 years of age (see Table 1). Additionally we see that at any given age, risk for men is approximately double that for women.

**Table 1:** COVID-19 deaths by age and sex per 100,000 population.

| Age Group | Sex | Population | COVID-19 Deaths | COVID-19 deaths per 100,000 population |
| --- | --- | --- | --- | --- |
| <55 | Male | 19,754,779 | 757 | 3.8 |
| <55 | Female | 19,395,980 | 445 | 2.3 |
| 55-59 | Male | 1,809,613 | 650 | 35.9 |
| 55-59 | Female | 1,861,038 | 301 | 16.2 |
| 60-69 | Male | 2,880,038 | 2,143 | 74.4 |
| 60-69 | Female | 3,028,537 | 1,044 | 34.5 |
| 70+ | Male | 3,383,401 | 13,984 | 413.3 |
| 70+ | Female | 4,173,575 | 11,280 | 270.3 |

Note: population data from ONS 2019 mid-year populations, COVID-19 deaths from ONS deaths between 1<sup>st</sup> March 2020 and 30<sup>th</sup> April 2020

The results of converting these mortality rates into risk scores and adding ethnicity specific adjustments to get Risk Scores A and B, in which BAME GPs have double or quadruple the risk of white GPs of the same age and sex respectively are shown in Table 2. Using either risk scoring system we find that GPs under 55 years of age regardless of sex or ethnicity are at the lowest risk whilst those over 70 years old are at the highest risk. We categorise GPs into four categories based on these risk scores: Low (0–1 points), Medium (1–4), High (4–9) and Very High (>9). Risk categories were defined to reflect discontinuities in the risk scores. Table 2 shows that the highest risk GPs in each of the risk categories is at approximately half the risk as the lowest risk GPs in the next risk category. The categorisation we have used works for both Risk Scores A and B and is robust to the choice of ethnicity adjustment.

**Table 2:** COVID-19 risk categorisation based on mortality risk.

| Age group | Sex | Ethnicity | Risk Score A<br>(BAME = 2*White) | Risk Score B<br>(BAME = 4*White) | COVID-19 Risk |
| --- | --- | --- | --- | --- | --- |
| 70+ | Male | BAME | 51.1 | 102.2 | very high |
| 70+ | Female | BAME | 33.4 | 66.8 | very high |
| 70+ | Male | White | 25.6 | 25.6 | very high |
| 60-69 | Male | BAME | 9.2 | 18.4 | very high |
| 70+ | Female | White | 16.7 | 16.7 | very high |
| 55-59 | Male | BAME | 4.4 | 8.9 | high |
| 60-69 | Female | BAME | 4.3 | 8.5 | high |
| 60-69 | Male | White | 4.6 | 4.6 | high |
| 55-59 | Female | BAME | 2 | 4 | medium |
| 55-59 | Male | White | 2.2 | 2.2 | medium |
| 60-69 | Female | White | 2.1 | 2.1 | medium |
| 55-59 | Female | White | 1 | 1 | low |
| <55 | Male | BAME | 0.5 | 0.9 | low |
| <55 | Female | BAME | 0.3 | 0.6 | low |
| <55 | Male | White | 0.2 | 0.2 | low |
| <55 | Female | White | 0.1 | 0.1 | low |

Notes: BAME = Black, Asian and Minority Ethnic, age-groups based on those used in the NHS risk reduction framework, GPs with missing data on age, sex or ethnicity classed with COVID-19 risk level Unknown

Summary statistics describing the 45,858 GPs and 639 single hander GP practices used in our analysis are given in Table 3. The vast majority of GPs (86.2%) and just over half of single-handed GP practices are classed as low risk. 4.9% of GPs and 14.9% of single-handed GP practices are classed as high risk and 3.0% of GPs and 17.8% of single-handed GP practices are classed as very high risk. Increased risk is particularly concentrated in single-handed practices almost 1/3 of which are run by a GP at high or very high risk from COVID.

**Table 3:**
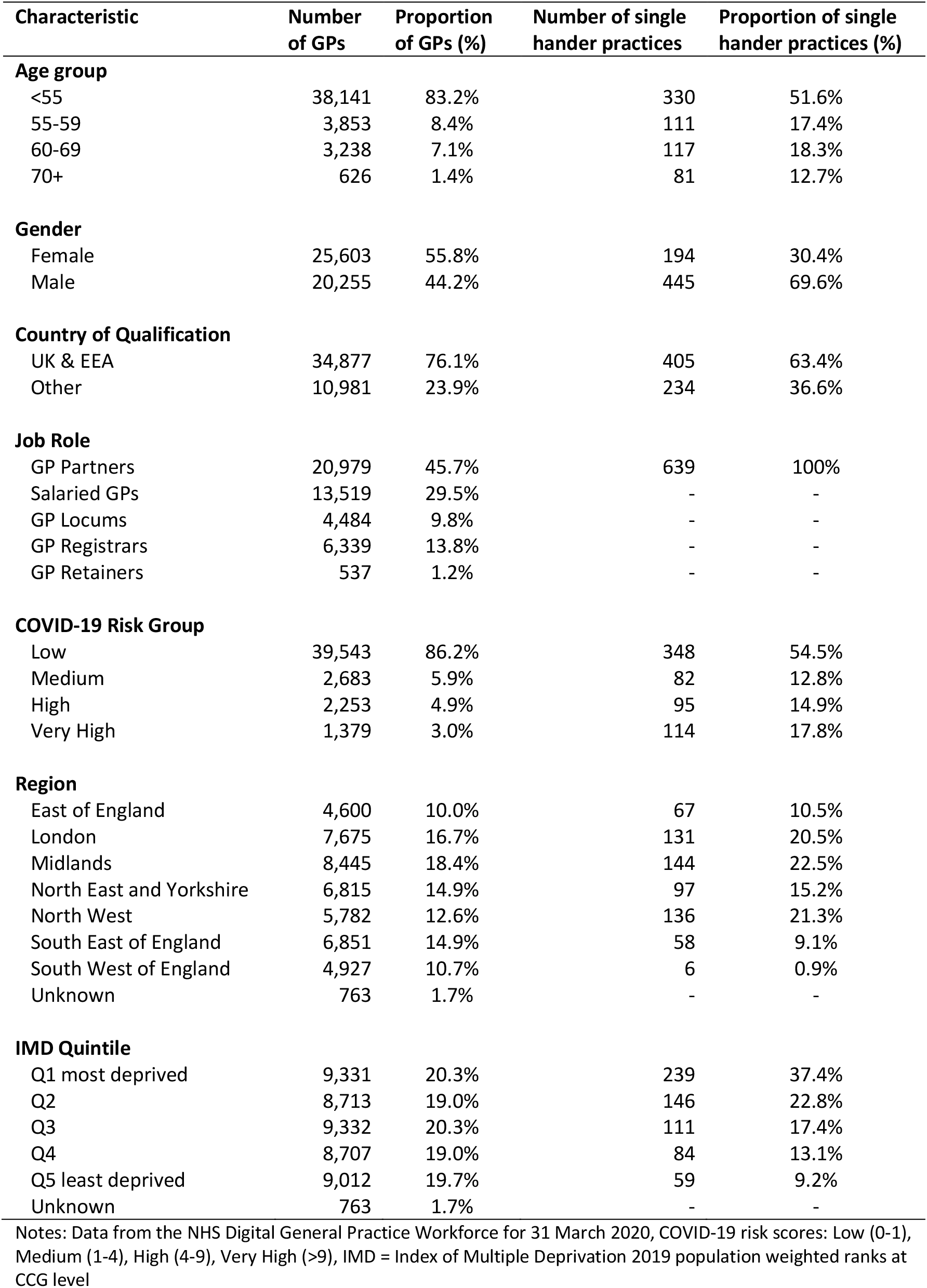
Descriptive statistics for the 45,858 GPs and 639 single hander GP practices in England.

Breaking down the GP workforce into age, sex and ethnicity groups (see Figure 1) shows that almost all GPs over the age of 70 are also of BAME ethnicity. We also see that locums are substantially over-represented amongst very high risk GPs, making up 17% of very high risk GPs (see Figure 1) whilst constituting less than 10% of the overall GP workforce (see Table 3). Full breakdowns of GPs by age, sex, ethnicity, risk group and job role are given in the supplementary appendix Table A2.

**Figure 1:**
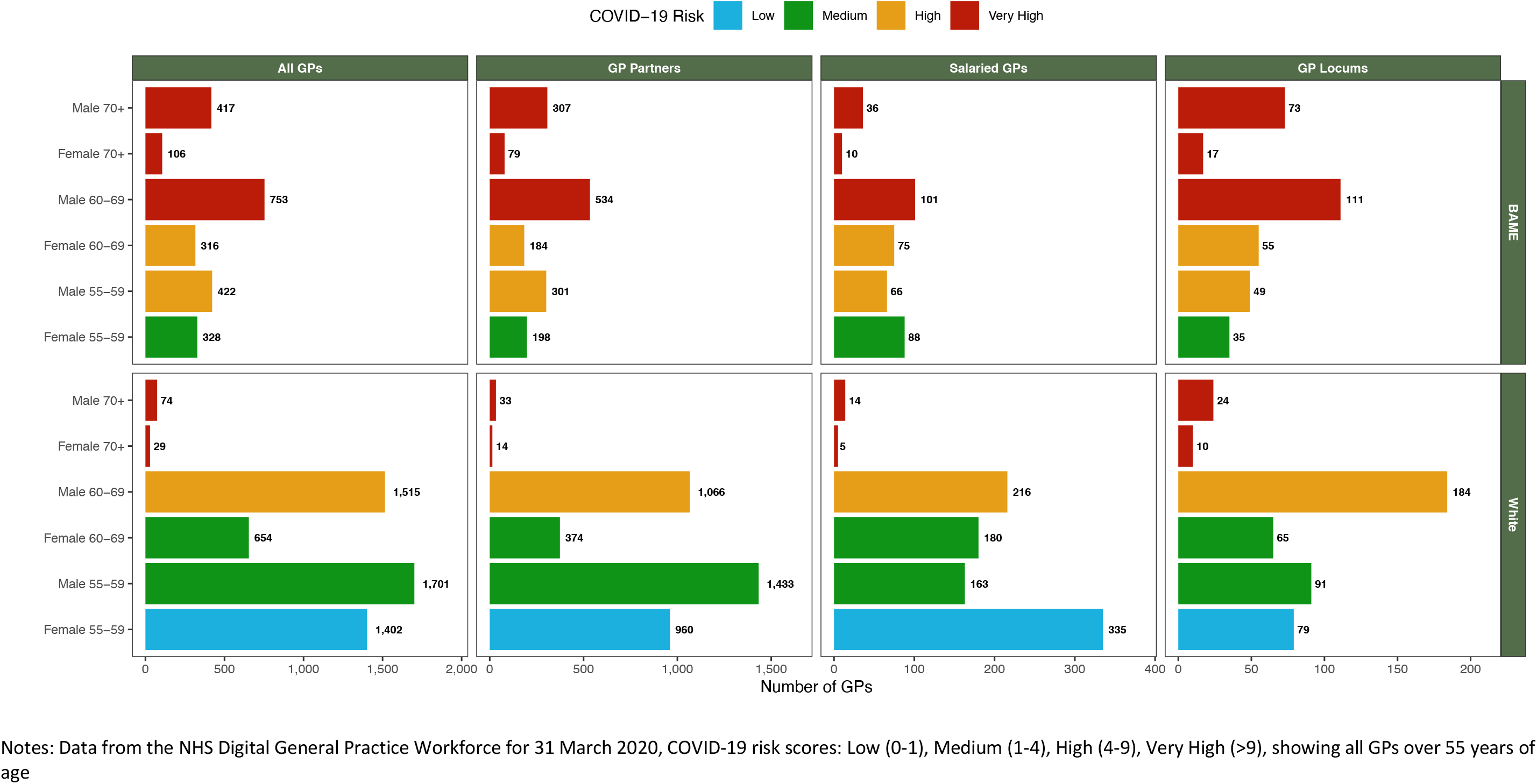
GP workforce breakdown by COVID-19 mortality risk

Examining the distribution of GPs according to the deprivation quintile of the CCG in which they practice (see Figure 2) we find that there is a steep socioeconomic gradient in the distribution of very high risk GPs. Very high risk GPs are more than 3 times as likely to be working in the most deprived CCGs in the country as they are to be working in the most affluent CCGs.

**Figure 2:**
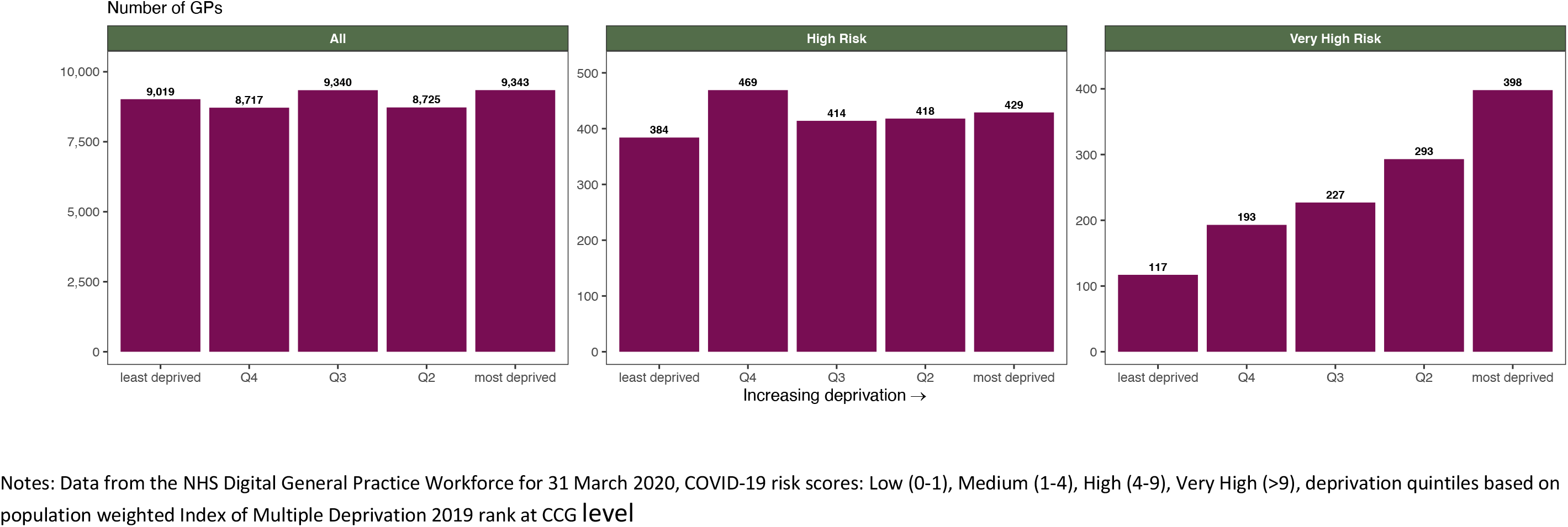
Socioeconomic distribution of GPs with high and very high risk of COVID-19 mortality

The socioeconomic distribution of single-handed GP practices displays an even steeper deprivation gradient (see Figure 3). Single-handed practices run by GPs classed as being at very high risk are more than 4 times as likely to be located in the most deprived CCGs in the country as compared to in the most affluent CCGs. We can also examine this through the numbers of patients registered to these single-handed practices (see Figure 4). There are 126,412 patients registered to single-handed GP practices classed as being at very high risk located in the most deprived CCGs in the country as compared to 33,745 patients registered to single-handed GP practices classed as very high risk located in the most affluent CCGs.

**Figure 3:**
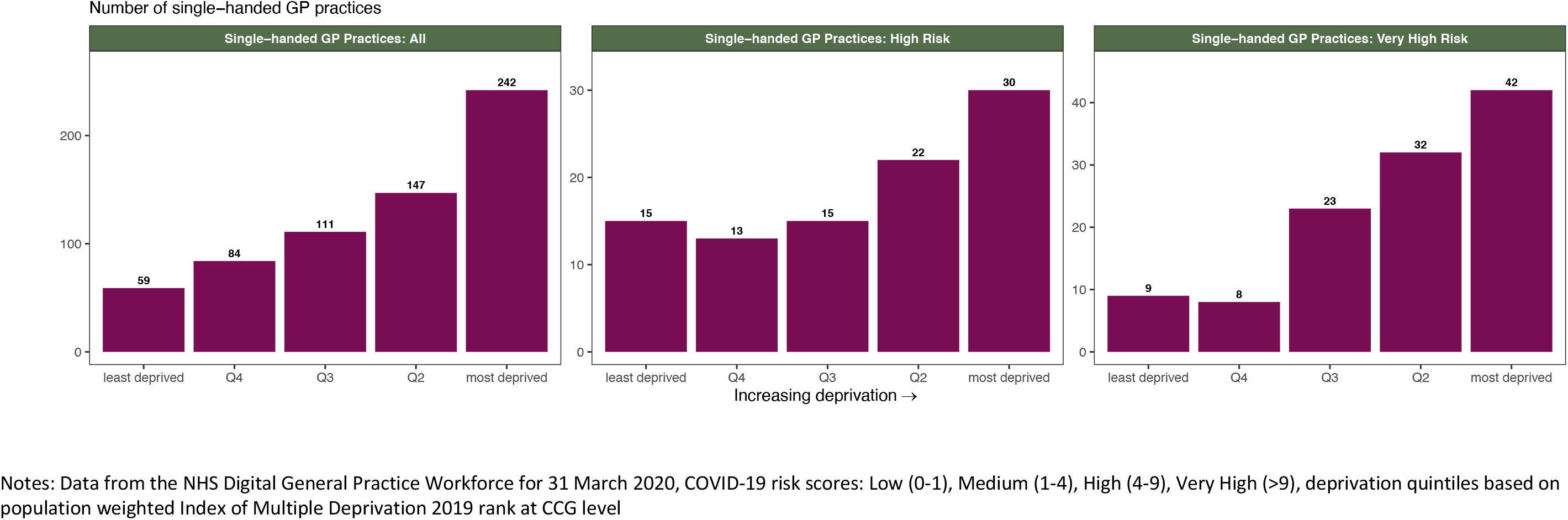
Socioeconomic distribution of single-handed GP practices run by GPs with high and very high COVID-19 mortality risk

**Figure 4:**
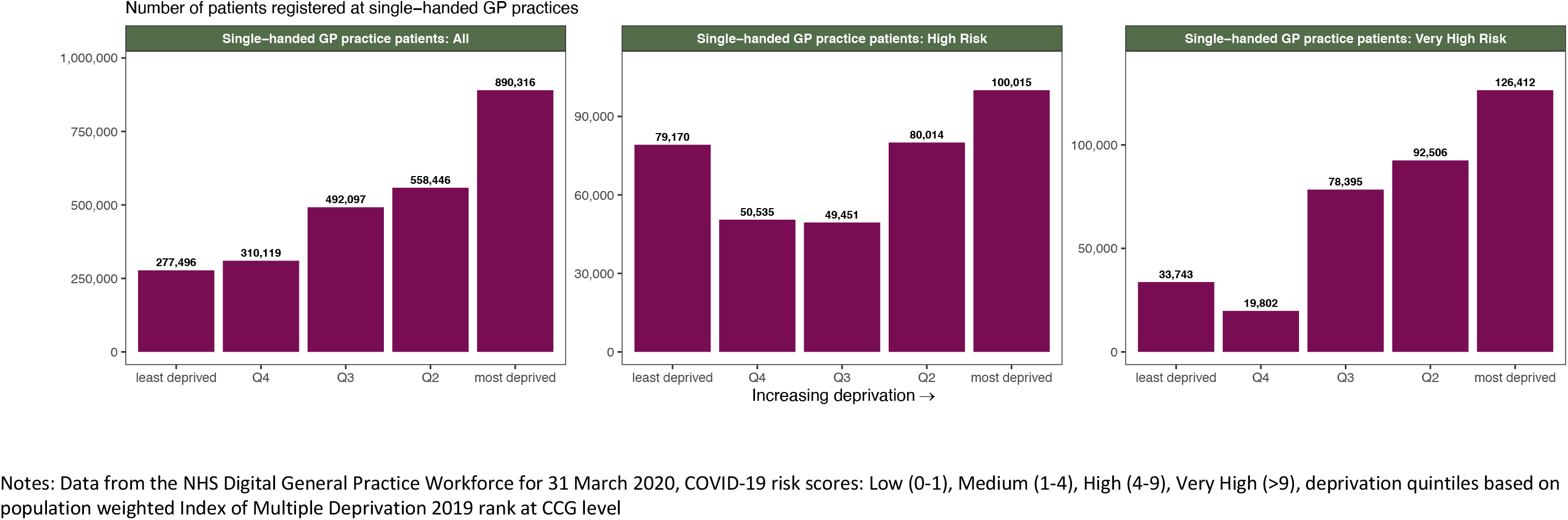
Socioeconomic distribution of patients registered at single-handed GP practices run by GPs with high and very high COVID-19 mortality risk

Examining the regional distribution of our three outcome measures (see Figure 5) we see that London has the highest proportions of very high risk GPs (5.2 very high risk GPs per 100,000 population), very high risk single-handed GP practices (0.37 very high risk single-handed GP practices per 100,000 population) and patients registered to very high risk single-handed GP practices (1,160 patients registered to very high risk single-handed GP practices per 100,000 population). Full breakdowns of our main outcomes by region are detailed in the supplementary appendix Table A6.

**Figure 5:**
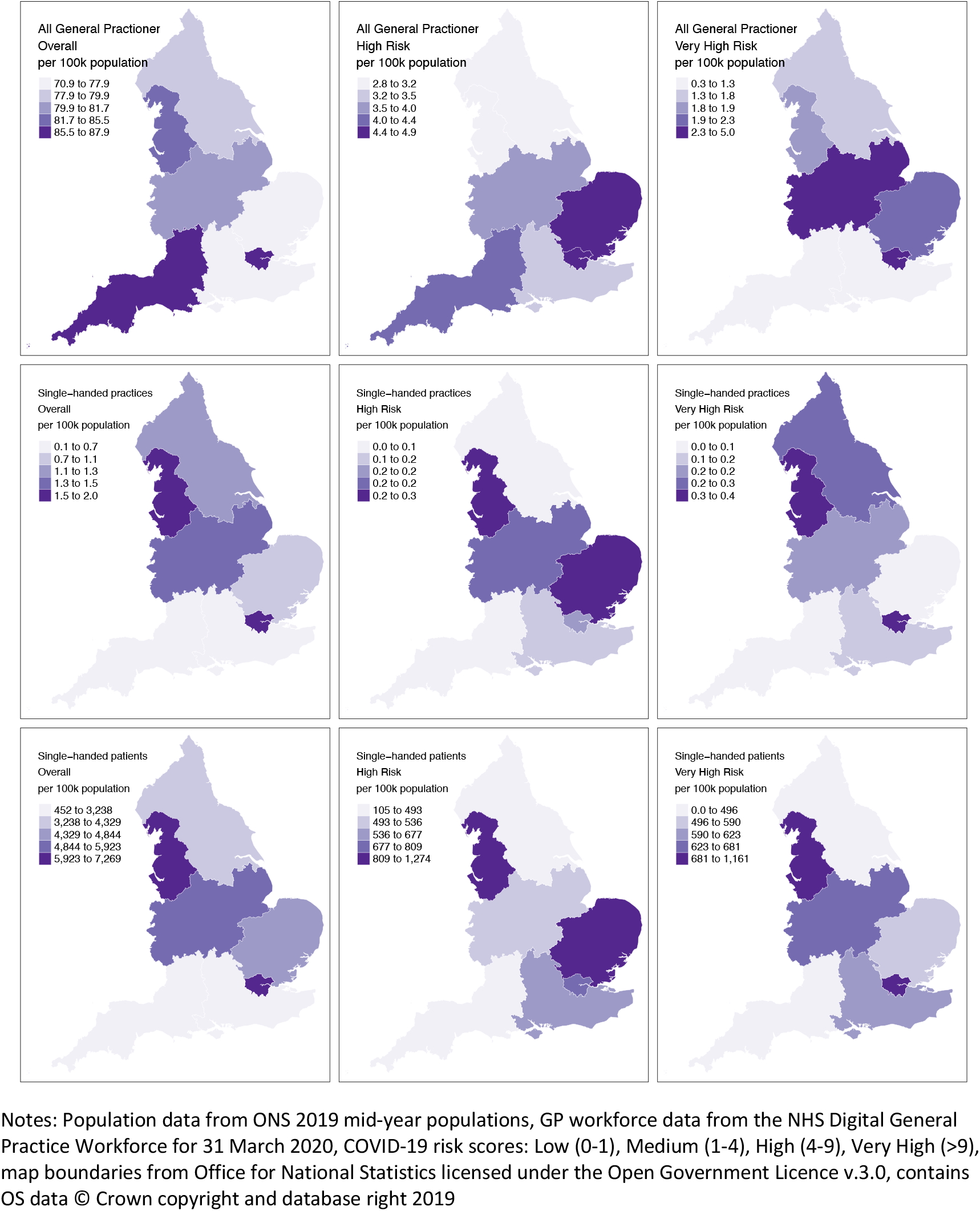
Regional distribution of GPs, single-handed practices and patients registered to these practices by COVID-19 mortality risk

## Discussion

### Statement of principle findings

Although the majority of GPs practicing in England are at low risk of death from COVID, a significant proportion of GPs, 7.9% (3,632 GPs) are at high or very high risk. These GPs are more likely to work in areas of high socioeconomic deprivation. Almost one in three single handed GP practices (32.7%) is run by a GP at high or very high risk from COVID. These single-handed GP practices are even more heavily concentrated in areas of high deprivation, particularly in London.

### Strengths and weaknesses

Our study is the first that we are aware of that explores the potential impact of COVID across the GP workforce in the NHS. We use a comprehensive national dataset to quantify the degree of fragility of primary care in the face of the COVID pandemic and highlight the particularly vulnerable position of single-handed GP practices in the delivery of primary care in these times. We also explore the implications of these risks on the regional and socioeconomic distributions of primary care provision and find that risks are patterned by both geography and deprivation. If left un-mitigated, existing health inequalities amongst the patient population are likely to be exacerbated along these dimensions.

Our study builds on emerging frameworks that identify COVID risk amongst healthcare staff in the NHS.(5,14) These frameworks were written to identify risk at the individual level, requiring detailed information about underlying health conditions and biomarkers of those being assessed. Our attempt to operationalise these frameworks at a health system level is challenging and has a number of limitations. First, detailed data on underlying health conditions, pregnancy, and biomarkers such as BMI and Vitamin D levels that are found in these frameworks are not recorded in the comprehensive national data sets that cover the NHS workforce underpinning our analysis. We were therefore unable to use these factors in our implementations of the COVID risk scores. Consideration of these additional factors if they were available would re-classify some GPs as being at higher risk than the risk level attributed to them in our analysis. Hence, the numbers of GPs we report to be at high COVID risk should be seen as conservative. Second, we were unable to obtain explicit data on ethnicity from the datasets used in our analysis. We instead had to use country of qualification as a proxy for ethnicity. This under-estimates the number of BAME people in our dataset assuming all GPs trained in the UK and EEA are White. The effect of this is likely to be a further under-estimation of the number of GPs at high risk from COVID. To get a sense of the magnitude of this bias we note that 23.9% of GPs in our dataset are classified as being of BAME ethnicity as compared to 44% of all doctors in the NHS.(18) We have requested bespoke data from NHS Digital and the GMC to explicitly capture ethnicity in our analysis and will update our results should this data become available to us.

Finally, we examined the socioeconomic distribution of high COVID risk GPs and single-handed GP practices using CCG level deprivation scores. CCGs cover large and heterogeneous populations and much of the granularity of the impacts of deprivation are masked when used at this coarse level of geography. Unfortunately, the datasets underpinning our analysis were unable to provide finer grained information on the geographical distribution of GPs and so we were limited by the data to perform our socioeconomic analysis at CCG level. We therefore expect that the socioeconomic gradients highlighted by our analysis will be much steeper in reality and could be better estimated if more granular geographic data on the distribution of GPs was available.

### Implications for policymakers

COVID creates numerous challenges for policy makers and commissioners, and amidst the inevitable focus on ‘re-opening’ secondary care, the implications of coexisting with COVID in general practice must not be overlooked. Maintaining general practice as a ‘front door’ to the NHS that is safe for both GPs and patients is vital but not easy. Options to quarantine and pre-test patients intended to help protect secondary care cannot be deployed in primary care. Precautions will be taken, but patient-facing members of the primary care team will be exposed to risk from COVID. Measures intended to protect higher risk GPs from COVID are likely to be necessary for some time, and may vary over time depending on COVID incidence and prevalence.

Withdrawing ‘high risk’ GPs from face to face consulting does not necessarily mean removing them from the clinical workforce. Doctors who are unable to see patients face to face may continue to consult via other means, including telephone and video consulting. Some ‘high risk’ GPs may decide to continue to see patients ‘as usual’. But surgeries need to plan for how to cover gaps in the provision of face to face appointments, acknowledging that the duration of GPs absence from face to face work is unknown. The scale of this challenge will vary depending on factors including the number of other GPs working at the same practice, and the COVID risk status of those GPs. Where a ‘single handed’ GP falls into a high risk group, practices may have no face to face provision at all. If GPs at higher risk do continue to practice face to face there is a greater than average risk that their ability to do so may be restricted by illness or death from COVID.

The impact of a reduction in GPs able to consult face to face is not evenly distributed geographically or by socioeconomic deprivation. This is likely to exacerbate inequalities in GP workload and funding which existed prior to COVID, and which result in GPs in areas of higher deprivation having a proportionately higher workload, with relative under-funding.(19–22) Our analysis shows a steep deprivation gradient. If GPs at higher risk of COVID stop seeing patients face to face, the reduction in provision will be greatest in the most deprived areas. These are the areas where overall health need is greatest, and where morbidity and mortality from COVID is likely to be greater too.(23,24) There are also concerns that alternatives to face to face consulting such as video consultations may be harder to access for deprived populations, and those with additional barriers to care such as English as a second language.

As GPs are likely to feel the weight of responsibility for their patients, local systems must work together – at primary care network and CCG level – to ensure that GPs are able to make the right personal choices for themselves around COVID risk, safe in the knowledge that patient care will not suffer. Increasing collaboration between surgeries in primary care networks may be one avenue for this, and exploring a locum market significantly changed by COVID is another.(25)

### Unanswered questions and future research

Analysis of occupational health risk from COVID should be expanded to include assessment of wider general practice teams, including allied health professionals and administrative support. Although the implications for face to face consulting may be different, risk assessment for the entire patient-facing practice staff must not be overlooked, particularly in light of evidence suggesting that COVID risk may be higher in less well paid roles.(26)

This study alerts us to a relatively large number of GPs at high risk of mortality from COVID, and to geographical and socioeconomic variation in the distribution of affected GPs. We do not know how many of these GPs will in practice choose to step away from direct patient contact, and how this may vary over time. Further work will be required to track what actually happens, and the effect on patient care of a possible reduction in the number of GPs able to consult face to face. There is a timely opportunity to provide additional support to mitigate COVID-19 risk for high risk GPs, GP practices and their patients. Failure to do so will likely further exacerbate existing health inequalities.

## Data Availability

Data used in the manuscript are all publicly available

## Acknowledgements

The authors would like to acknowledge helpful comments from Health Foundation colleagues.

## Data sharing statement

All data used in this study is publicly available administrative data. The data sources have been highlighted in the methods section of the manuscript. Code for all the analyses as well as the anonymised database will be made available on reasonable request.

## Supplementary appendix

**Table A1:** Population, IMD 2019 score and IMD 2019 quintile for Clinical Commissioning Groups as of April 2020

| CCG Code | CCG Name<br>(from April 2020) | Population<br>(mid-2018) | IMD 2019<br>Score | IMD 2019<br>Quintile |
| --- | --- | --- | --- | --- |
| 00R | NHS Blackpool CCG | 139,305 | 45.39 | 1 |
| 01J | NHS Knowsley CCG | 149,571 | 43.14 | 1 |
| 99A | NHS Liverpool CCG | 494,814 | 42.44 | 1 |
| 03F | NHS Hull CCG | 260,645 | 40.58 | 1 |
| 14L | NHS Manchester CCG | 547,627 | 39.87 | 1 |
| 05L | NHS Sandwell and West Birmingham CCG | 504,641 | 37.78 | 1 |
| 00Q | NHS Blackburn with Darwen CCG | 148,942 | 36.15 | 1 |
| 01D | NHS Heywood, Middleton and Rochdale CCG | 220,001 | 34.67 | 1 |
| 01G | NHS Salford CCG | 254,408 | 34.32 | 1 |
| 05W | NHS Stoke on Trent CCG | 264,149 | 33.99 | 1 |
| 00Y | NHS Oldham CCG | 235,623 | 33.69 | 1 |
| 15E | NHS Birmingham and Solihull CCG | 1,179,020 | 33.62 | 1 |
| 36J | NHS Bradford District and Craven CCG | 587,936 | 32.93 | 1 |
| 01T | NHS South Sefton CCG | 159,237 | 32.79 | 1 |
| 07L | NHS Barking and Dagenham CCG | 211,998 | 32.74 | 1 |
| 06A | NHS Wolverhampton CCG | 262,008 | 32.41 | 1 |
| 01F | NHS Halton CCG | 128,432 | 32.34 | 1 |
| 05Y | NHS Walsall CCG | 283,378 | 32.07 | 1 |
| 07T | NHS City and Hackney CCG | 288,371 | 32.03 | 1 |
| 01X | NHS St Helens CCG | 180,049 | 31.69 | 1 |
| 00N | NHS South Tyneside CCG | 150,265 | 31.68 | 1 |
| 03H | NHS North East Lincolnshire CCG | 159,821 | 31.34 | 1 |
| 00T | NHS Bolton CCG | 285,372 | 31.03 | 1 |
| 16C | NHS Tees Valley CCG | 674,284 | 30.96 | 1 |
| 04C | NHS Leicester City CCG | 355,218 | 30.84 | 1 |
| 00P | NHS Sunderland CCG | 277,417 | 30.70 | 1 |
| 02X | NHS Doncaster CCG | 310,542 | 30.46 | 1 |
| 02P | NHS Barnsley CCG | 245,199 | 29.99 | 1 |
| 01Y | NHS Tameside and Glossop CCG | 258,613 | 29.75 | 1 |
| 12F | NHS Wirral CCG | 323,235 | 29.73 | 1 |
| 01A | NHS East Lancashire CCG | 380,013 | 29.63 | 1 |
| 03L | NHS Rotherham CCG | 264,671 | 29.63 | 1 |
| 08M | NHS Newham CCG | 352,005 | 29.49 | 1 |
| 13T | NHS Newcastle Gateshead CCG | 502,704 | 29.28 | 1 |
| 03J | NHS North Kirklees CCG | 192,750 | 28.88 | 1 |
| 08V | NHS Tower Hamlets CCG | 317,705 | 27.63 | 2 |
| 15F | NHS Leeds CCG | 789,194 | 27.58 | 2 |
| 03R | NHS Wakefield CCG | 345,038 | 27.36 | 2 |
| 03N | NHS Sheffield CCG | 582,506 | 27.25 | 2 |
| 10R | NHS Portsmouth CCG | 215,133 | 27.20 | 2 |
| 10X | NHS Southampton CCG | 252,796 | 26.88 | 2 |
| 84H | NHS County Durham CCG | 526,980 | 26.61 | 2 |
| 02T | NHS Calderdale CCG | 210,082 | 26.46 | 2 |
| 06P | NHS Luton CCG | 214,109 | 26.05 | 2 |
| 02H | NHS Wigan Borough CCG | 326,088 | 25.89 | 2 |
| 07P | NHS Brent CCG | 330,795 | 25.66 | 2 |
| 08W | NHS Waltham Forest CCG | 276,700 | 25.24 | 2 |
| 05X | NHS Telford and Wrekin CCG | 177,799 | 24.99 | 2 |
| 05C | NHS Dudley CCG | 320,626 | 24.31 | 2 |
| 01E | NHS Greater Preston CCG | 202,562 | 24.07 | 2 |
| 00V | NHS Bury CCG | 190,108 | 23.92 | 2 |
| 52R | NHS Nottingham and Nottinghamshire CCG | 1,037,356 | 23.68 | 2 |
| 10L | NHS Isle of Wight CCG | 141,538 | 23.33 | 2 |
| 05A | NHS Coventry and Rugby CCG | 473,979 | 23.27 | 2 |
| 11N | NHS Kernow CCG | 568,210 | 23.01 | 2 |
| 93C | NHS North Central London CCG | 1,498,001 | 22.97 | 2 |
| 07W | NHS Ealing CCG | 341,982 | 22.75 | 2 |
| 02Q | NHS Bassetlaw CCG | 116,839 | 22.75 | 2 |
| 06T | NHS North East Essex CCG | 338,326 | 22.68 | 2 |
| 03A | NHS Greater Huddersfield CCG | 245,977 | 22.56 | 2 |
| 99G | NHS Southend CCG | 182,463 | 22.55 | 2 |
| 08C | NHS Hammersmith and Fulham CCG | 185,426 | 22.33 | 2 |
| 08Y | NHS West London CCG | 226,099 | 22.28 | 2 |
| 72Q | NHS South East London CCG | 1,811,249 | 22.25 | 3 |
| 03K | NHS North Lincolnshire CCG | 172,005 | 22.22 | 3 |
| 99C | NHS North Tyneside CCG | 205,985 | 22.22 | 3 |
| 00L | NHS Northumberland CCG | 320,274 | 22.08 | 3 |
| 05H | NHS Warwickshire North CCG | 193,752 | 21.80 | 3 |
| 01H | NHS North Cumbria CCG | 318,631 | 21.79 | 3 |
| 01K | NHS Morecambe Bay CCG | 330,572 | 21.72 | 3 |
| 26A | NHS Norfolk & Waveney CCG | 1,021,991 | 21.72 | 3 |
| 07Y | NHS Hounslow CCG | 270,782 | 21.57 | 3 |
| 07G | NHS Thurrock CCG | 172,525 | 21.08 | 3 |
| 01W | NHS Stockport CCG | 291,775 | 20.99 | 3 |
| 09D | NHS Brighton and Hove CCG | 290,395 | 20.86 | 3 |
| 15M | NHS Derby and Derbyshire CCG | 1,019,900 | 20.51 | 3 |
| 91Q | NHS Kent and Medway CCG | 1,846,478 | 20.36 | 3 |
| 71E | NHS Lincolnshire CCG | 755,833 | 20.27 | 3 |
| 15N | NHS Devon CCG | 1,194,166 | 20.17 | 3 |
| 97R | NHS East Sussex CCG | 554,590 | 19.80 | 3 |
| 15C | NHS Bristol, North Somerset and South Gloucestershire CCG | 959,968 | 19.72 | 3 |
| 01V | NHS Southport and Formby CCG | 116,159 | 19.51 | 4 |
| 99E | NHS Basildon and Brentwood CCG | 262,412 | 19.49 | 4 |
| 04Y | NHS Cannock Chase CCG | 136,974 | 19.28 | 4 |
| 02E | NHS Warrington CCG | 209,547 | 19.05 | 4 |
| 78H | NHS Northamptonshire CCG | 730,650 | 18.78 | 4 |
| 02M | NHS Fylde and Wyre CCG | 192,421 | 18.66 | 4 |
| 02G | NHS West Lancashire CCG | 113,949 | 18.61 | 4 |
| 11X | NHS Somerset CCG | 559,399 | 18.60 | 4 |
| 09A | NHS Central London (Westminster) CCG | 185,422 | 18.55 | 4 |
| 08G | NHS Hillingdon CCG | 304,824 | 18.36 | 4 |
| 18C | NHS Herefordshire and Worcestershire CCG | 784,164 | 18.32 | 4 |
| 05D | NHS East Staffordshire CCG | 128,418 | 18.25 | 4 |
| 04F | NHS Milton Keynes CCG | 275,002 | 17.77 | 4 |
| 06L | NHS Ipswich and East Suffolk CCG | 409,248 | 17.72 | 4 |
| 00X | NHS Chorley and South Ribble CCG | 176,862 | 17.70 | 4 |
| 05G | NHS North Staffordshire CCG | 219,571 | 17.62 | 4 |
| 08N | NHS Redbridge CCG | 303,858 | 17.30 | 4 |
| 05N | NHS Shropshire CCG | 320,274 | 17.13 | 4 |
| 11J | NHS Dorset CCG | 772,268 | 17.05 | 4 |
| 08F | NHS Havering CCG | 257,810 | 17.03 | 4 |
| 10V | NHS South Eastern Hampshire CCG | 216,379 | 16.94 | 4 |
| 06H | NHS Cambridgeshire and Peterborough CCG | 889,112 | 16.85 | 4 |
| 27D | NHS Cheshire CCG | 721,292 | 16.21 | 4 |
| 02A | NHS Trafford CCG | 236,370 | 16.13 | 4 |
| 02Y | NHS East Riding of Yorkshire CCG | 317,404 | 16.09 | 4 |
| 07K | NHS West Suffolk CCG | 230,997 | 15.99 | 4 |
| 36L | NHS South West London CCG | 1,494,905 | 15.99 | 4 |
| 42D | NHS North Yorkshire CCG | 426,821 | 15.65 | 4 |
| 05Q | NHS South East Staffordshire and Seisdon Peninsula CCG | 226,137 | 15.52 | 5 |
| 07H | NHS West Essex CCG | 306,910 | 15.16 | 5 |
| 08E | NHS Harrow CCG | 250,149 | 15.10 | 5 |
| 11M | NHS Gloucestershire CCG | 633,558 | 14.94 | 5 |
| 06F | NHS Bedfordshire CCG | 455,229 | 14.75 | 5 |
| 70F | NHS West Sussex CCG | 858,852 | 14.44 | 5 |
| 92G | NHS Bath and North East Somerset, Swindon and Wiltshire CCG | 918,428 | 14.28 | 5 |
| 15D | NHS East Berkshire CCG | 434,463 | 13.99 | 5 |
| 10K | NHS Fareham and Gosport CCG | 201,622 | 13.96 | 5 |
| 06K | NHS East and North Hertfordshire CCG | 569,078 | 13.80 | 5 |
| 99F | NHS Castle Point and Rochford CCG | 177,051 | 13.70 | 5 |
| 06Q | NHS Mid Essex CCG | 393,065 | 13.56 | 5 |
| 05V | NHS Stafford and Surrounds CCG | 155,803 | 13.55 | 5 |
| 04V | NHS West Leicestershire CCG | 402,165 | 13.29 | 5 |
| 10J | NHS North Hampshire CCG | 223,795 | 12.12 | 5 |
| 03Q | NHS Vale of York CCG | 362,955 | 11.96 | 5 |
| 06N | NHS Herts Valleys CCG | 595,670 | 11.94 | 5 |
| 05R | NHS South Warwickshire CCG | 270,064 | 11.89 | 5 |
| 15A | NHS Berkshire West CCG | 489,709 | 11.80 | 5 |
| 10Q | NHS Oxfordshire CCG | 672,414 | 11.72 | 5 |
| 11A | NHS West Hampshire CCG | 566,879 | 11.37 | 5 |
| 03W | NHS East Leicestershire and Rutland CCG | 335,800 | 10.78 | 5 |
| 99M | NHS North East Hampshire and Farnham CCG | 211,590 | 10.40 | 5 |
| 92A | NHS Surrey Heartlands CCG | 1,036,742 | 10.32 | 5 |
| 14Y | NHS Buckinghamshire CCG | 542,512 | 10.15 | 5 |
| 10C | NHS Surrey Heath CCG | 96,474 | 9.24 | 5 |

**Table A2:** Distribution of COVID-19 mortality risk by age, sex and ethnicity – data underpinning figure 1

| Age Group | Sex | Ethnicity | COVID-19 mortality risk | All GPs | GP Partners | Salaried GPs | GP Locums | GP Registrars | GP Retainers |
| --- | --- | --- | --- | --- | --- | --- | --- | --- | --- |
| 70+ | Female | BAME | Very High | 106<br>(0.23%) | 79<br>(0.17%) | 10<br>(0.02%) | 17<br>(0.04%) | 0<br>(0%) | 0<br>(0%) |
| 70+ | Female | White | Very High | 29<br>(0.06%) | 14<br>(0.03%) | 5<br>(0.01%) | 10<br>(0.02%) | 0<br>(0%) | 0<br>(0%) |
| 70+ | Male | BAME | Very High | 417<br>(0.91%) | 307<br>(0.67%) | 36<br>(0.08%) | 73<br>(0.16%) | 0<br>(0%) | 1<br>(0%) |
| 70+ | Male | White | Very High | 74<br>(0.16%) | 33<br>(0.07%) | 14<br>(0.03%) | 24<br>(0.05%) | 0<br>(0%) | 3<br>(0.01%) |
| 60-69 | Male | BAME | Very High | 753<br>(1.64%) | 534<br>(1.16%) | 101<br>(0.22%) | 111<br>(0.24%) | 3<br>(0.01%) | 4<br>(0.01%) |
| 60-69 | Female | BAME | High | 316<br>(0.69%) | 184<br>(0.4%) | 75<br>(0.16%) | 55<br>(0.12%) | 0<br>(0%) | 2<br>(0%) |
| 60-69 | Male | White | High | 1,515<br>(3.3%) | 1,066<br>(2.32%) | 216<br>(0.47%) | 184<br>(0.4%) | 1<br>(0%) | 48<br>(0.1%) |
| 55-59 | Male | BAME | High | 422<br>(0.92%) | 301<br>(0.66%) | 66<br>(0.14%) | 49<br>(0.11%) | 5<br>(0.01%) | 1<br>(0%) |
| 60-69 | Female | White | Medium | 654<br>(1.43%) | 374<br>(0.82%) | 180<br>(0.39%) | 65<br>(0.14%) | 0<br>(0%) | 35<br>(0.08%) |
| 55-59 | Female | BAME | Medium | 328<br>(0.72%) | 198<br>(0.43%) | 88<br>(0.19%) | 35<br>(0.08%) | 2<br>(0%) | 5<br>(0.01%) |
| 55-59 | Male | White | Medium | 1,701<br>(3.71%) | 1,433<br>(3.12%) | 163<br>(0.36%) | 91<br>(0.2%) | 0<br>(0%) | 14<br>(0.03%) |
| 55-59 | Female | White | Low | 1,402<br>(3.06%) | 960<br>(2.09%) | 335<br>(0.73%) | 79<br>(0.17%) | 0<br>(0%) | 28<br>(0.06%) |
| <55 | Female | BAME | Low | 4,694<br>(10.24%) | 1,407<br>(3.07%) | 1,771<br>(3.86%) | 622<br>(1.36%) | 850<br>(1.85%) | 44<br>(0.1%) |
| <55 | Female | White | Low | 18,074<br>(39.41%) | 6,109<br>(13.32%) | 7,316<br>(15.95%) | 1,307<br>(2.85%) | 3,012<br>(6.57%) | 330<br>(0.72%) |
| <55 | Male | BAME | Low | 3,945<br>(8.6%) | 1,797<br>(3.92%) | 821<br>(1.79%) | 590<br>(1.29%) | 735<br>(1.6%) | 2<br>(0%) |
| <55 | Male | White | Low | 11,428<br>(24.92%) | 6,183<br>(13.48%) | 2,322<br>(5.06%) | 1,172<br>(2.56%) | 1,731<br>(3.77%) | 20<br>(0.04%) |

**Table A3:** Distribution of COVID-19 mortality risk for GPs by CCG IMD quintile – data underpinning figure 2

| IMD 2019 Quintile | All GPs | High Risk GPs | Very High Risk GPs |
| --- | --- | --- | --- |
| Q5 least deprived | 9,012 | 414 | 143 |
| Q4 | 8,707 | 488 | 209 |
| Q3 | 9,332 | 464 | 290 |
| Q2 | 8,713 | 437 | 312 |
| Q1 most deprived | 9,331 | 450 | 425 |
| Unknown | 763 | 0 | 0 |

**Table A4:** Distribution of COVID-19 mortality risk for single handed GP practices by IMD quintile – data underpinning figure 3

| IMD 2019 Quintile | All | High Risk | Very High Risk |
| --- | --- | --- | --- |
| Q5 least deprived | 59 | 15 | 9 |
| Q4 | 84 | 13 | 8 |
| Q3 | 111 | 15 | 23 |
| Q2 | 146 | 22 | 32 |
| Q1 most deprived | 239 | 30 | 42 |

**Table A5:** Distribution of COVID-19 mortality risk for patients registered to single handed GP practices by IMD quintile – data underpinning figure 4

| IMD 2019 Quintile | All | High Risk | Very High Risk |
| --- | --- | --- | --- |
| Q5 least deprived | 277,496 | 79,170 | 33,743 |
| Q4 | 310,119 | 50,535 | 19,802 |
| Q3 | 492,097 | 49,451 | 78,395 |
| Q2 | 546,183 | 80,014 | 92,506 |
| Q1 most deprived | 871,264 | 100,015 | 126,412 |

**Table A6:** Rates of GPs, single-handed GP practices, and patients registered to single-handed GP practices per 100,000 population

| Region | All General Practitioners Overall | All General Practitioners High Risk | All General Practitioners Very High Risk | Single-handed practices Overall | Single-handed practices High Risk | Single-handed practices Very High Risk | Single-handed patients Overall | Single-handed patients High Risk | Single-handed patients Very High Risk |
| --- | --- | --- | --- | --- | --- | --- | --- | --- | --- |
| East of England | 70.84 | 4.53 | 2.11 | 1.03 | 0.28 | 0.14 | 4,684 | 1,273 | 580 |
| London | 86.16 | 5.00 | 5.22 | 1.47 | 0.17 | 0.37 | 6,028 | 741 | 1,160 |
| Midlands | 80.14 | 3.90 | 2.47 | 1.37 | 0.19 | 0.20 | 4,950 | 505 | 634 |
| North East and Yorkshire | 79.55 | 3.08 | 1.95 | 1.13 | 0.14 | 0.21 | 4,092 | 489 | 475 |
| North West | 82.45 | 3.44 | 2.10 | 1.94 | 0.21 | 0.29 | 6,996 | 826 | 692 |
| South East | 77.39 | 4.08 | 2.03 | 0.66 | 0.15 | 0.15 | 3,023 | 581 | 605 |
| South West | 87.89 | 4.23 | 0.41 | 0.11 | 0.04 | - | 451 | 105 | - |

